# Estimation of SARS-CoV-2 fitness gains from genomic surveillance data without prior lineage classification

**DOI:** 10.1101/2024.01.08.24300976

**Authors:** Tjibbe Donker, Alexis Papathanassopoulos, Hiren Ghosh, Raisa Kociurzynski, Marius Felder, Hajo Grundmann, Sandra Reuter

**Affiliations:** Institute for Infection Prevention and Control. Medical Center University of Freiburg, Faculty of Medicine, University of Freiburg, Germany

## Abstract

The emergence of SARS-CoV-2 variants with increased fitness has had a strong impact on the epidemiology of COVID-19, with the higher effective reproduction number of the viral variants leading to new epidemic waves. Tracking such variants and their genetic signatures, using data collected through genomic surveillance, is therefore crucial for forecasting likely surges in incidence. Current methods of estimating fitness advantages of variants rely on tracking the changing proportion of a particular lineage over time, but describing successful lineages in a rapidly evolving viral population is a difficult task. We propose a new method of estimating fitness gains directly from nucleotide information generated by genomic surveillance, without a-priori assigning isolates to lineages from phylogenies, based solely on the abundance of Single Nucleotide Polymorphisms (SNPs). The method is based on mapping changes in the genetic population structure over time. Changes in the abundance of SNPs associated with periods of increasing fitness allow for the unbiased discovery of new variants, and thereby obviating a deliberate lineage assignment and phylogenetic inference. We conclude that the method provides a fast and reliable way to estimate fitness advantages of variants without the need for a-priori assigning isolates to lineages.

## Introduction

Since its emergence in late 2019, SARS-CoV-2 has caused billions of infections worldwide [2]. Ever since, successive epidemic waves have repeatedly devastated population health, exhausted public health care systems and eroded the economic bases of societies in an unprecedented manner [8, 15]. Whilst some of these waves may have been driven by seasonal variations, changes in risk behaviour among populations, or a relaxation in control measures imposed by governments, so far, most pandemic waves were rekindled by the emergence of viral variants with higher transmissibility or the capacity of evading existing immunity in humans, or a combination of these phenotypes [11]. The threat of emerging variants has prompted WHO to classify certain SARS-CoV-2 lineages as Variants of Concern (VOC), Variants of Interest (VOI) or Variants under Monitoring (VUM)[3, 13]. Yet, the classification depends on a risk assessment by the Technical Advisory Group on SARS-CoV-2 Virus Evolution (TAG-VE). This takes into consideration the abundance and expansion of named lineages established by the scientific nomenclature system such as those used by Nextstrain [1] and Pango-lineage [20], as well as their phenotypic and public health impact. Expansion of a lineage leading to an increasing relative prevalence is an indication of improved fitness of the emerging variant, causing an increase in the effective reproduction number (*R*_*e*_) of the combined viral population [16]. The increasing relative prevalence is a direct measure of the relative fitness i.e. the competitive advantage over the pre-existing viral population which will be replaced and excluded from the host population, eventually fuelling another incipient wave. Many of the current methods of estimating fitness advantage rely on this increasing prevalence of a particular lineage over time [19, 4]. Such methods thus require a division of the viral population into lineages with each being tested for potential fitness advantages.

Decisions on the classification of variants or lineages are non trivial and hardly unequivocal considering the size of evolving viral populations and the ubiquitous emergence of diversity during an unfolding pandemic, resulting in the constant addition of Single Nucleotide Polymorphisms (SNP) to the ever expanding phylogeny. Designating each isolate containing a novel SNP (or any novel combinations thereof) as a variant is justified in a strict sense, but would defy the purpose of lineage classification as it would not make taxonomic sense as long as its progeny has not reached a certain level of abundance. As a consequence, the question of what defines a lineage, requires more information about the abundance and diversity of underlying variant [21]. However, even then, it remains unclear if the chosen division is meaningful in terms of the viral ecology (i.e. fitness).

As a result, a multitude of variants would need to be tested for potential fitness gains, including any combination of closely related lineages combined, in order to discriminate emerging lineages in a meaningful way. Moreover, telling apart abundance changes that occur as a result of random genetic drift from those that occur on the basis of true fitness advantage threatens any deliberate classifications of VUMs, VOIs, and VOCs.

We here propose an alternative attempt for estimating relative fitness directly from the SNP data, obviating time-consuming and compute-intensive phylogenetic reconstruction and the need of dividing the viral population into variants or lineages. Additionally, using available sequencing data directly from nucleotide archives should improve speed and efficiency of the entire process when millions of isolates are collected and sequenced.

We show how changes in the fitness of the virus can be directly estimated from genomic surveillance data, without the a-priori need to define lineages. The presented methods focus on the viral population as a whole, and track its changes in genetic composition. This is achieved by quantifying and tracking the prevalence of extant SNPs in the sequence collection. Changes in the relative abundance indicate shifts in the viral population, and are a marker of the emergence of potential variants of concern. Isolates belonging to potential variants underlying these movements are then identified post-hoc, based on how the SNPs they contain developed over time resulting in a more natural delineation of variants of concern.

## Results

We set out to estimate the fitness advantage of a potential variant within the viral population using data from genomic surveillance. We downloaded the genomic data from GISAID [22] in the form of a multi-sequence alignment (MSA) of all submitted sequences until 11 October 2022, and selected all sequences from Germany (Available at gisaid.org/EPI SET 240108qz). In total, the dataset included 797,206 sequences, between 18-01-2020 and 11-10-2022, with a notable increase in the number of sequences per week from 05-03-2021 (week 10, 2021) onwards (Figure S1, Section S2).We track all Single Nucleotide Polymorphisms (SNPs), defined as any position within the MSA that differs from the reference sequence hCoV-19/Wuhan/WIV04/2019 (EPI-ISL-402124), irrespective of their abundance. We include all insertions present in the MSA as SNPs, bringing the total number of potential positions to 32561 (29891 bases in the reference genome and 2670 insertions present in the included isolates), with SNPs present at 86% (N=27933) of the positions. There were 51614 unique combinations of base and positions (i.e. unique SNPs), a large proportion of the SNPs (n=9067, 18%) is present in only a single sequence, while only few (n=30, 0.004%) are present in more than 50% of the sequences (Figure S2).

We start from the theoretical case where a single mutation causes a nucleotide change at a single position in the genome (i.e. a single SNP). This single SNP increases the reproductive success of the virus, expressed as its net growth rate in terms of newly infected individuals. The mechanism (increased infectiousness, increased infectious duration, immune evasion, etc.[4]) is not important in this respect, as long as the growth rate increases. As the virus containing this fitness increasing SNP spreads faster, the part of the viral population at a given time *t* containing the SNP (*p*_*t*_) grows according to a predictable trajectory, described as a sigmoidal function of the added fitness advantage.

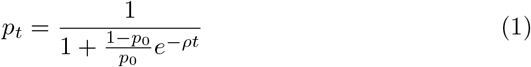

where *ρ* is the added fitness advantage in terms of the net growth rate difference between the wildtype and the variant.

We can use this to estimate the fitness advantage of this single SNP if we have reliable observations of its proportion over time from genomic surveillance, because the difference in logit of these proportions is a linear function of the fitness advantage

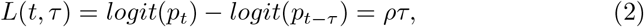

and therefore

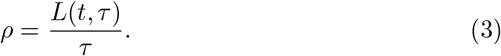

where *τ* is the time between the current time *t* and compared time *t−τ* . The observed SNP proportions follow the same sigmoidal function as projected based on the fitness estimates (figure 1A), although some SNPs either do not start or end at a proportion near 0 or 1, causing differences in the fitness estimates (See supplementary section S3).

**Figure 1:**
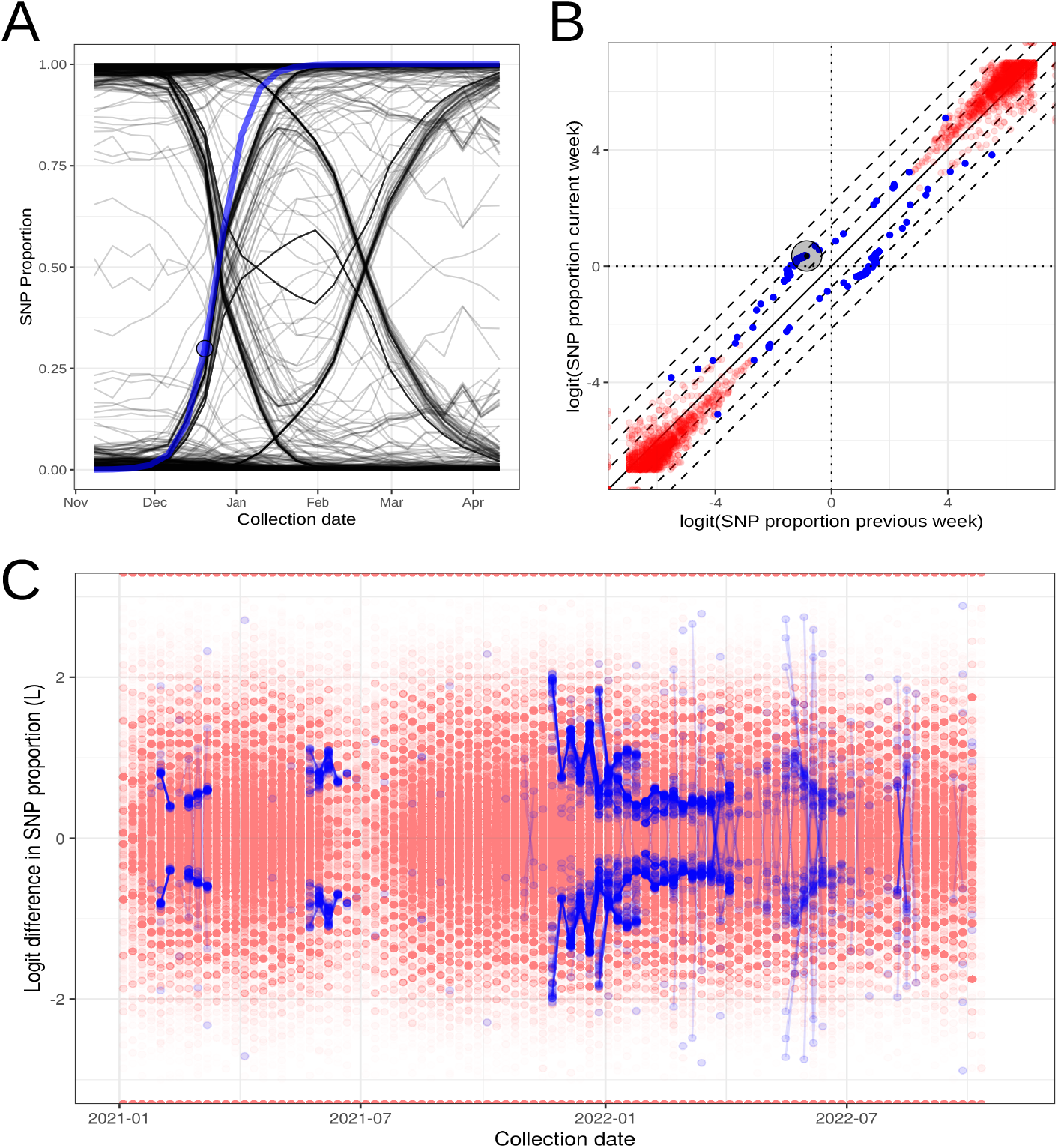
Direct fitness advantage estimates per SNP. A) Each line shows the proportion of a single SNP over time, the blue line shows the projected proportion of the example SNP in the obsercation week (circle) based on the assumption that difference logit(p) remains the same as in the observation week. B) The logit of the SNP proportion for week 1 of 2022 and the previous week for each SNP. Blue dots show included SNPs with significant distance from the diagonal, red dots show excluded SNPs. Larger black circle shows the example SNP. C) The logit(p) difference for each SNP in each week (dots), compared to the previous week. Significantly different SNPs are connected between successive weeks.

We can then estimate the fitness advantage of any SNP present in two time frames, e.g. two consecutive weeks (figure 1B), by taking the logit difference in proportion in both time frames. This can also be done by visually inspecting the logit phase plane (plotting logit proportion of one week on the x-axis and the other week on the y-axis). Although we observe clear groups of SNPs with the same apparent fitness advantage, most SNPs remain in the bottom-left or top-right corner of the graph, corresponding to very low or very high proportions in both weeks, where near 0 or 1 proportions cause large variations in the fitness advantage estimate. The low proportion SNPs can be filtered out (figure 1B red dots), based on the confidence interval on the estimate, delivering a clearer estimate but at the expense of potential early detection. Even the SNPs included after filtering (figure 1B blue dots) show high variability in fitness estimates over time (figure 1C), resulting in a highly variable median estimated fitness advantage.

Instead of estimating the fitness advantage of single SNPs, we can also use the entire genetic population structure of the virus to get to a single estimate of fitness advantage. We propose doing this by measuring the genetic divergence, analogous to *F*_*ST*_ [9], between the viral population in a given reference week and each individual preceding and successive week. While such metrics are usually used to measure the divergence of contemporaneous populations in different locations, it can in principle also measure the amount the current population diverges from past populations in the same location. If the population is under strong selective pressure, or if more fit individuals are introduced, the genetic population structure can rapidly change.

We do this by measuring the genetic diversity within the combined population of both time periods (*H*_*s*_), as defined by Donker et al [6], relative to the genetic diversity between the two populations (*H*_*p*_). This change is reflected in a diversity measurement between the current (*t*) and a past or future (*t* + *τ*) population. If a variant with sufficient fitness advantage is growing within the viral population, the difference in 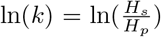 will follow an approximately linear function away from the reference week with each successively compared week (figure 2A). Any genetic variation in the viral population causing SNPs to be partially present in the wildtype or variant will cause this decline in ln(*k*) to be slower, thus slightly underestimating the fitness advantage (figure 2B). We therefore obtain the best estimate of the current fitness changes (figure 2C&D) if we consider the maximum weekly ln(*k*) difference in recent weeks (2 weeks ≤*τ*≤ 8 weeks).

**Figure 2:**
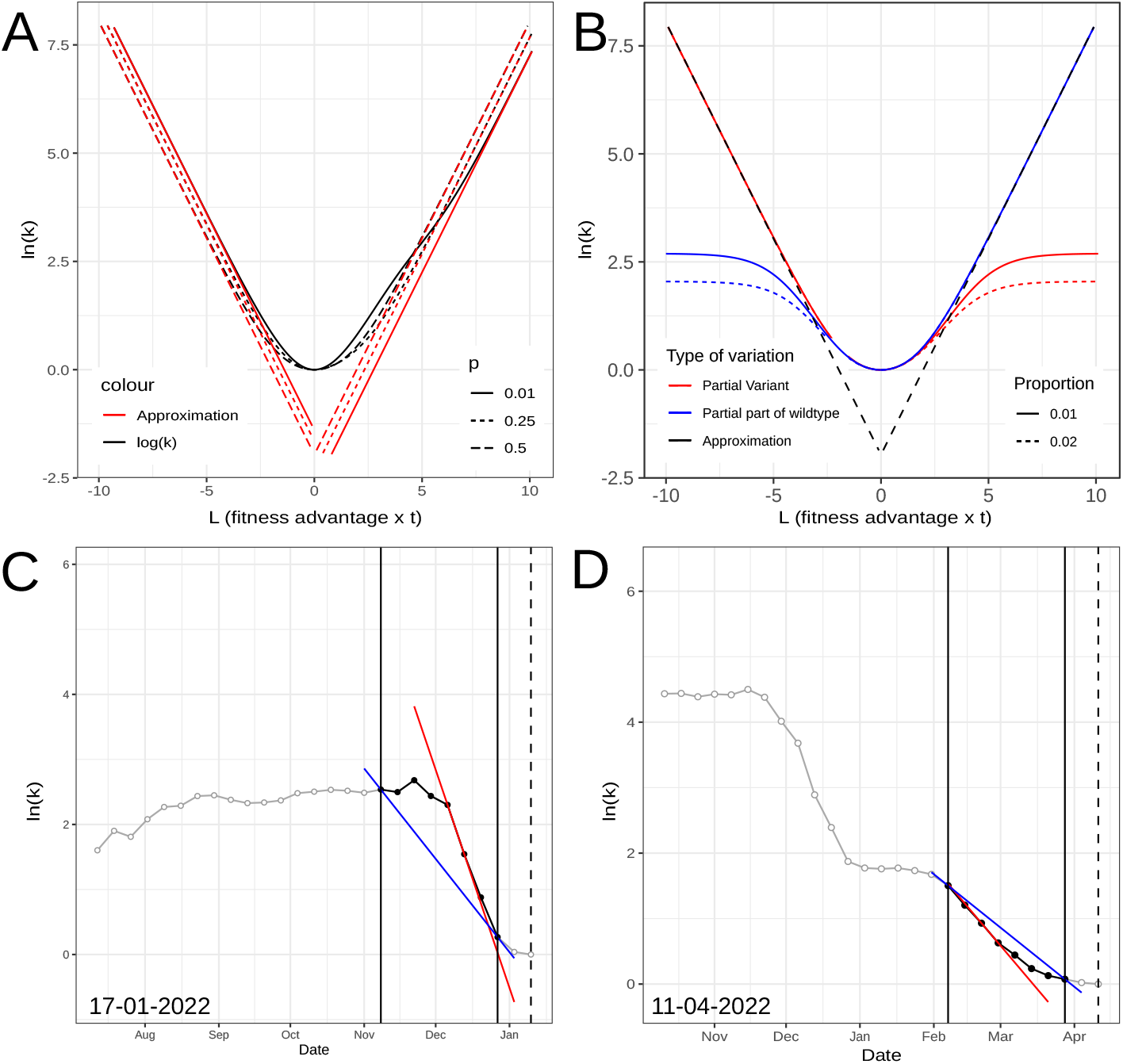
Fitness advantage estimate based on the change in viral population divergence. A) The expected trajectory of ln(*k*(*t*)) and the approximation Δ ln(*k*(*t*)) ≈*abs*(*L*(*t*)) for SNPs unique to the variant, and B) SNPs partially present in the variant (red) or partially present in the wildtype (blue). C&D) Examples of measured ln(*k*) over time, showing the mean (blue) and maximum (red) slope within the measuring timeframe.

By successively taking each week as reference week, we then obtain an estimate of the variant fitness changes over time (figure 3A). We can clearly observe the clusters of equal fitness related to the emergence of the Delta, Omicron, Omicron BA.2, and Omicron BA.5 variants (figure 3B). The emergence of Omicron BA.1 and BA.2 falls within the same period of increased fitness, probably because BA-2 followed quickly on from BA.1. The arrival of the Alpha variant is also visible, albeit to a lesser extent due to the lower number of isolates underlying the earlier weeks in the data. For each of the known variants, the method detected the effect of the variant’s increased fitness on the genetic population structure before the effective reproduction number *R*_*e*_ increases (figure 3C), and well before the incidence increases (figure 3D). We observe a significant correlation between the ln(*k*) value and *R*_*e*_ (Figure S3), particularly when we consider the *R*_*e*_ two weeks in the future (Figure S3C),indicating that *R*_*e*_ tends to rise after the method detects a fitness gain.

**Figure 3:**
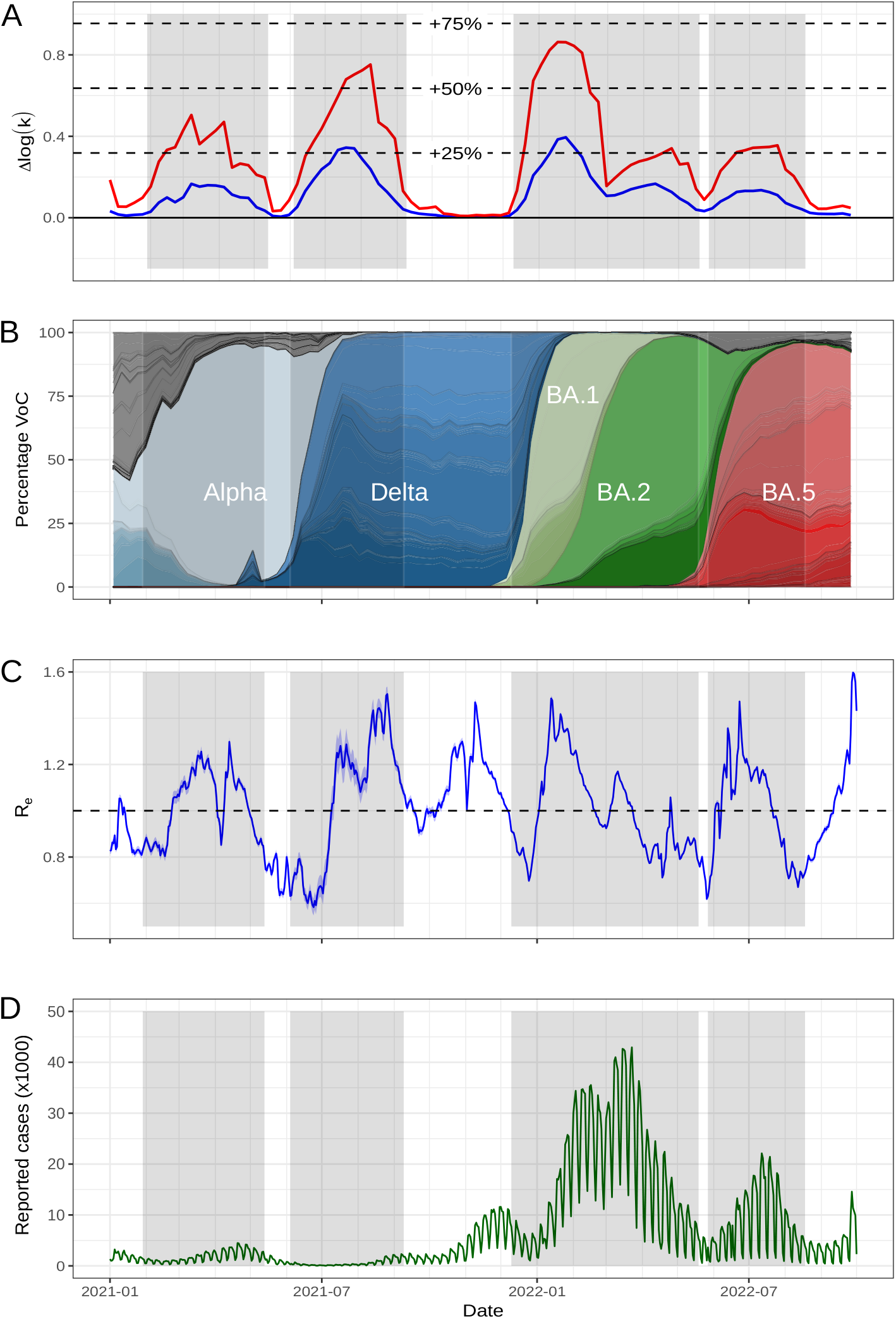
The viral population-based estimate. A) Fitness advantage estimate based on the logit ln(*k*) difference, the blue line shows the median of the estimates, the red line shows the maximum estimates. Shaded areas show the times when the estimated fitness advantage was above 12.5%. B) The proportion of the isolates belonging to each pango-lineage per week. C & D) Effective reproduction number and reported number of cases, respectively, for the same time period.

The estimated fitness advantages for the Alpha, Delta, BA.1/BA.2, and BA.5 time periods, respectively +40%, +59%, +68%, and +28%, based on the maximum slope of ln(*k*) were generally in line with estimates by others [23, 7, 24], with the exception of the Omicron BA.1 advantage over Delta. The estimated BA.1 fitness advantage for Germany was estimated lower than other estimates, which usually estimate it above +100%. Estimates based on sequences collected in other countries yield higher estimates for BA.1 (UK: +126%, USA: +130%, South Africa: +147%, Figure S4), while estimates from a single state in Germany result in the same fitness gain estimates (Figure S5).These differences in estimates for BA.1 can potentially be explained by differences in immunity in the host population between these countries, as BA.1’s fitness advantage is primarily caused by its ability to evade pre-existing immunity.

The same periods of fitness differences are found when using fewer sequences; by selecting as few as 100 random isolates from each week, we get a consistently similar result in terms of both the fitness increase time frames and the estimated fitness advantage (Figure S6). Selecting even fewer isolates (10 to 50 per week) results in higher variation in the ln(*k*) estimates, making detection less reliable. Using the periods of largest fitness differences, we can select the isolates that likely belong to the variant, based on the combined fitness advantage estimates of the SNPs they contain (i.e. their sequence total SNP advantage score). We observe a correct overlap with the known variants (figure 4, tables S3-S7). In one time period (between 26 December 2021 and 14 May 2022) the score indicates that the vast majority of isolates are part of the variant (Table S5). However, these isolates include both Omicron BA.1 and BA.2 variants, which on closer inspection (figure 4C) fall in two distinct groups with different total SNP scores.

**Figure 4:**
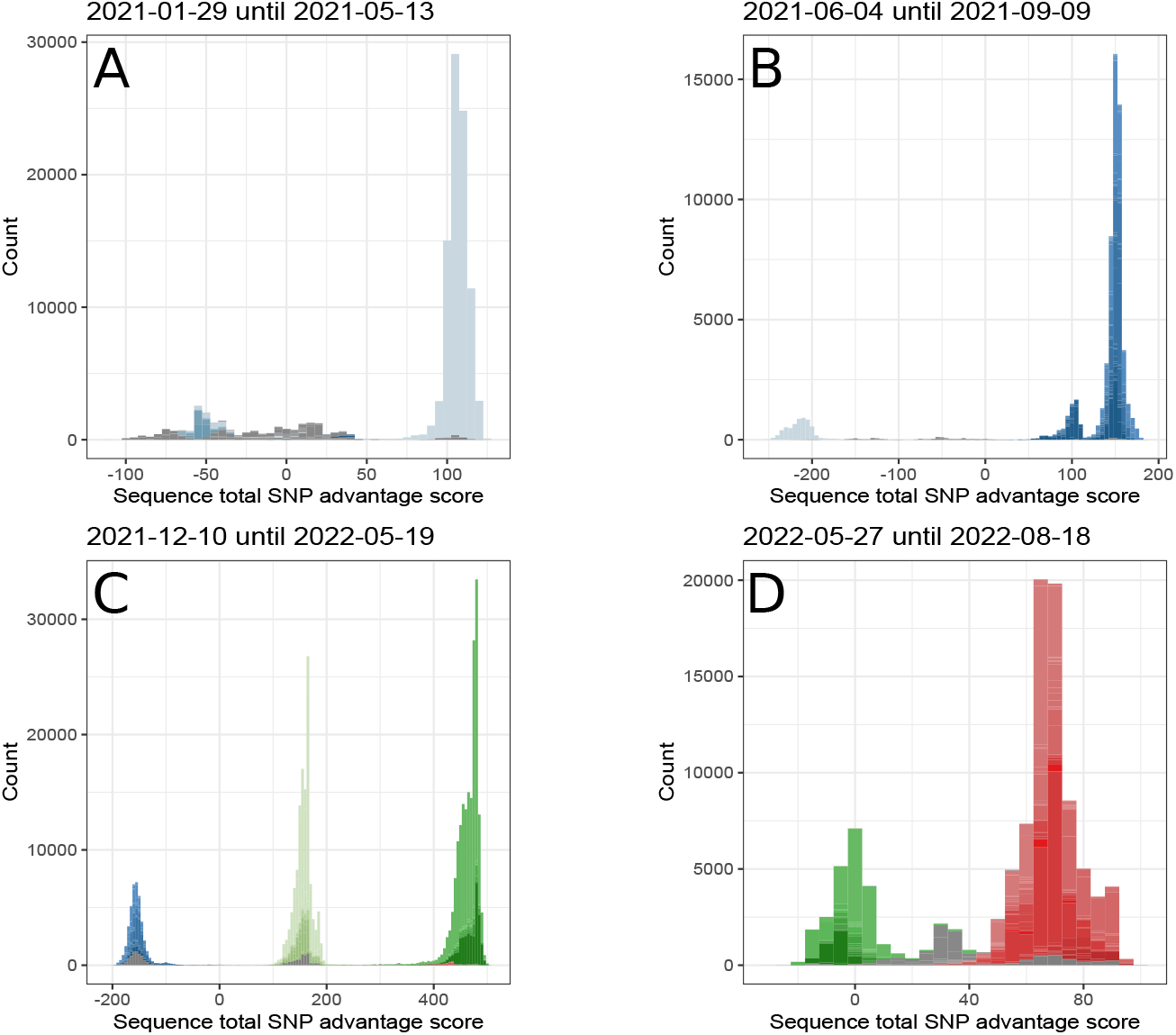
The total SNP advantage score per sequence during each of the four time frames during which the growth of a variant with increased fitness advantage was detected. Colours denote Pango lineages, grouped by the six major variants: Alpha (light blue), Delta (dark blue), Omicron BA.1 (light green), BA.2 (dark green), and BA.5 (red).

The BA.1 isolates still have a positive SNP score because most of their included SNPs keep increasing over the time period in question, because they are also included in the BA.2 isolates. The BA.2 isolates subsequently score higher because they include more increasing SNPs.

The fitness gain is generally detected early during the growth of the variant, with Delta at 9.8% of the week’s isolates at the moment of detection, BA.1 at 3.4%, and BA.5 at 13.9%. Only Alpha was detected at a considerably higher proportion (54.8%), which is likely caused by fewer available isolates which were likely not a representative sample at that time. When only considering the sequences that were submitted until the reference week, we observe that the fitness gains are detected about two weeks later (21-06-2021 vs 07-06-2021 for Delta, 27-12-2021 vs 13-12-2021 for BA.1 and 13-06-2022 vs 30-05-2022 for BA.5). This is primarily caused by lack of sequences submitted close to their isolation date, causing too much variation when using the current or previous week as reference (Figure S7). Additionally, the first detection of each variant is not influenced by the time window used for the Δ ln(*k*) calculation (Figure S8), although a comparison to viral populations long ago generally delivers higher estimates.

Furthermore, the method scales well with the number of samples included: the calculation time increased with the number of included sequences, but decreased per included sample (Figure S9). The method took 60.4 seconds to produce an estimate for Germany (N=797,206), 82.7 seconds for the UK (N=2,775,418 sequences), 128 seconds for the US (N=3,769,411), 38.8 seconds for Denmark (N=576,296), and 10.5 seconds for South Africa (N=35,770) on our benchmark desktop computer (see supplementary information Section S6).

## Discussion

We demonstrate that an agnostic mathematical approach for estimating fitness gains based on available genomic surveillance data can be a fast and reliable method for the identification of successful phenotypes without a-priori classification of variants and lineages. The presented methods deliver a robust fitness advantage estimate direct from the sequencing data, where conventional approaches rely not only on variant and lineage classification but also on phylogenetic inference. Whilst conventional methods rely on operational taxonomical units such as sequence types, phylogroups, and clades[5, 11] to impose a degree of order on an ever evolving population, we analyse the viral population in its entirety, relying on the signal left in the genetic structure of the pathogen population by the shift towards a more fit genotype.

This method analyses the viral population in its entirety, tracking the speed at which the complete population’s genetic composition changes. This is done by comparing the change over time in the genetic diversity within the combined population of both time points relative to the diversity between the two populations. In a way, any successful genotype (or variant) changes the genetic composition of the viral population at a predictable pace for a given fitness advantage. As we can measure the pace, we can determine the the fitness gain. Thus the method can detect perturbations in the genetic population structure, quantify fitness gains, and identify successful genomes whilst being agnostic of deliberate classification efforts. It therefore avoids any potential erroneous understanding of what a meaningful subdivision of the population is, instead dividing the isolates according to how they align with the estimated shift in fitness.

The fitness gains are estimated based on changes in proportion presence of all SNPs over time, irrespective of the effect each individual SNP has on the fitness of the virus. All SNPs unique to the variant, even the neutral ones, will therefore improve the estimator’s signal over noise ratio. Since the estimation is based on the rate of change in SNP proportions, the final estimate of fitness gain is independent of the number of included SNPs. The estimation is, however, influenced by genetic variation in the variant or wildtype population. If a SNP is partially present in the variant population, it will exhibit a slower growth than the variant itself, particularly towards a higher prevalence of the variant, thus causing an underestimate of the fitness gain. Similarly, if a SNP is present in the variant as well as partially in the wildtype, the initial growth and fitness of the variant will be underestimated. However, information from SNPs unique to the variant will aid the method towards an accurate estimate. Even if the SNPs conferring the actual fitness advantage are not unique to the variant, for instance because two SNPs with only a joint effect on fitness come together through recombination, the neutral SNPs unique to the original recombined genome will still allow for a reliable estimate.

The method scales well and is fast enough to potentially analyse all isolates collected globally. However, it implicitly assumes that both infected and susceptible hosts are randomly mixing, or at least that the replacement of the wildtype by the new variant is not hindered by the population structure of the hosts. This assumption still more or less holds when considering single countries or smaller geographical areas, but is not applicable when considering the global viral population. Globally, the replacement of variants will likely be influenced by the interplay between the dissemination of lineages across different geographical areas and the proliferation of the lineages within these areas. The time it takes the variant to disseminate globally will slow the replacement of the wildtype, resulting in a lower estimate of fitness change based on genetic composition of the global viral population. We therefore argue that these estimates are best done on the smallest geographical unit with enough sequences available.

The fact that the presented method does not require phylogenetic reconstruction to identify the variant and estimate its fitness advantage, gives it a major advantage over existing methods, since the construction of phylogenetic trees is challenging when millions of isolates are included. Although a number of methods have been designed to speed up this process, for instance by constructing a phylogenetic backbone of the tree based on a subset of the data and placing the rest of the sequences onto this backbone, we show that such a step can be left out all together. Furthermore, splitting of a phylogenetic tree brings with it the question of where the splits need to be placed. Splitting the tree too coarse will cause the variant to remain hidden among the wild-type for too long, while splitting it too fine will result in the variant itself being split up in many different lineages. Siding with caution, most lineage classification systems have a rather fine resolution, with multiple lineages making up the variants [1]. Because lineage classification is not straight-forward it often requires human input, leaving a lot of room for different interpretations and discussion. The presented method, on the other hand, gives consistently the same estimate for a given dataset, with only the threshold of what to call a variant (i.e. how high does the signal need to be to sound a variant alert) in human hands.

Although we produce estimates based on a large collection of assembled and aligned sequences, it should be possible to get similar estimates from samples containing a mixture of lineages, such as those resulting from wastewater surveillance. In such cases, the changes in the proportion presence of SNPs within samples already provides information on the fitness gains of certain lineages. Such proportions can be readily calculated as the number of reads containing the SNP mapped to a particular position divided by the total number of reads mapped to the same position [14]. This could potentially provide an earlier detection of fitness gains in the viral population, although this would require further study to assess its performance.

The used metric is analogous to Wright’s F statistic (*F*_*ST*_), as it compares genetic diversity within and between populations, but differs in a number of key aspects. In the most commonly used implementation of *F*_*ST*_ [18], diversity is defined as the population’s heterozygosity; the chance of randomly picking two individuals with different SNPs, from either the combined population or from the two population that are being compared. We, however, define diversity as the chance of picking the same pair of discordant SNPs from these populations, as previously defined [6]. This has the advantage that it is less influenced by large numbers of SNPs that are nearly fully present, or almost absent. Furthermore, we use ln(*k*) = ln(*H*_*s*_/*H*_*p*_) as the primary metric, because its theoretical range of (0 *…*∞), instead of (− ∞*…*∞) for *x* = *logit*(*F*_*ST*_), makes it less prone to extreme values when populations are genetically more similar, and makes recent measurements more reliable.

While tracking the changes in the viral genetic population structure delivers a reliable estimate of the underlying variant’s fitness advantage, it is unable to directly identify which isolates belong to the variant or the wildtype. To do so, we determined for each isolate if the SNPs contained in its genome moved in the direction of the fitness increase. In this process, as within the previous, we use information from all SNPs, and do not select SNPs on the basis of their position within the genome. Not all of the SNPs moving in the direction of the variant may thus cause a fitness advantage. There are most likely a lot of spurious SNPs and synonymous mutations that were part of the parental lineage the variant emergent from (i.e. hitch-hiking mutations) or that arose shortly after emergence of the variant. However, even the hitch-hiking mutations provide information on the changes in genetic population structure due to a disseminating variant with increased fitness, and can therefore also be used to determine whether or not an isolate belongs to the variant population. The method performs exceptionally well, as seen by the overlap with the pango lineage assignment of the samples. It should be noted that the presented methods are not a lineage classification system, and are not meant to be so, as they are designed to detect increases in fitness, which is only one part of what determines a lineage.

A preliminary estimate of fitness advantage of a potential variant can also be produced without calculating ln(*k*) by inspecting the logit proportion phase planes over time. This method of tracking individual SNP proportions works as long as it is not obscured by spurious large estimates at the very low or high SNP proportions. When a true variant is spreading, a small burst of SNPs should follow a line parallel to the diagonal in subsequent logit phase planes. The distance of this line to the diagonal then corresponds to the likely fitness advantage of the variant. However, it should be noted that the SNPs might not follow the exact same trajectory, as random variation within both the wild-type and variant population can affect these trajectories. While tracking the changes in the population prevalence of individual SNPs provides a relatively easy way of estimating the fitness advantage of the isolates containing the SNPs of interest, it does not deliver a reliable combined estimate for a single variant spreading through the population. For this, the genetic population-based method needs to be used.

## Limitations

The primary use of fitness estimations is to inform policy makers as well as modellers of impending increases of the effective reproduction number (*R*_*e*_) of the circulating disease. This means, however, that the variant and its fitness advantage need to be detected well before *R*_*e*_ starts to rise and a new potential wave starts. Although variants were detected before their respective epidemic waves, this does not mean that this result would be obtained when performing this analysis in real-time, because of delays between isolation, genome sequencing, and submission. In the current retrospective analysis, the data are treated like the sequences were submitted immediately after isolation. As our sensitivity analysis suggests, a live estimation on this dataset would probably have resulted in delayed detection by two weeks caused by the delay between isolation of the sample and submission of the sequence. However, this problem also applies to methods that rely on prevalences of an a-priori designated variant within the submitted sequencing data, reiterating the need for accelerating the process between isolation of the sample and the publication of the sequence in the database.

More generally, it should be noted that these methods rely on large, well-structured, genomic surveillance efforts. This became possible during the COVID-19 pandemic, when its added value became apparent after the identification of the Alpha variant in the UK. Although the concept of such genomic surveillance has become a mainstream idea in pandemic response planning, the availability of funding and infrastructure for the long term is far from certain in many countries. With fewer samples available, the estimates start to fluctuate, as visible in the first period of the study. However, we show that the algorithm produces a relatively stable estimate with about 100 sequences per week. Nonetheless, if such large genomic surveillance efforts are not maintained, methods like these will also be rendered less effective.

## Conclusion

We were successful in estimating the fitness advantage of SARS-CoV-2 variants directly from the genomic surveillance data, without the need for phylogenetic reconstruction. The presented methods are fast, scale well with the number of included isolates, and are independent of lineage classification. Most importantly, we show that the fitness advantage of a particular variant can be estimated without knowledge about which isolates belong to the variant or not, by studying the effect the variant has on the temporal-genetic structure of the entire viral population.

## Materials and Methods

The central aim of this study is to estimate the fitness advantage of an unknown variant in a viral population from genomic surveillance data. In general, the data should consist of assembled and aligned sequences of a particular virus with metadata on the date of isolation. We assume that the fitness advantage is genetically determined through point mutation, although the methods should also work for large insertion of genetic code, recombination, etc.

We refer to the viral population that existed prior to the emergence of the variant as the *wildtype* viral population, even if this population itself was the result of a previous variant establishing itself as the majority variant. We assume that all viruses in the wildtype viral population are equally fit. The fitness advantage of the variant (*ρ*) is defined as the increase in its growth rate relative to the wildtype growth rate.

## Data

We downloaded all available sequences on GISAID [22] as part of a multisequence alignment (MSA) on 1 November 2022. The dataset consisted of 12,331,461 full-length genomes with a length greater than 29,000 bp and less than 5% undetermined nucleotides (NNNNs), submitted until 11 October 2022. MSA was performed by MAFFT [12] as described on the GISAID platform. We selected all isolates from Germany, accessible at gisaid.org/EPI SET 240108qz (DOI: 10.55876/gis8.240108qz). We then calculated the proportion of each base *b* at that position *i* at time *t* (in weeks) *t* (*p*_*i,b,t*_) for all positions in the MSA that contained differences compared to the reference strain (hCoV-19/Wuhan/WIV04/2019, accession number EPI IS 402124), as the number of sequences containing base *b* at position *i*, divided by the number of sequences with a known base at that position. As the majority of these single nucleotide polymorphisms (SNPs) contain two majority bases, while the other two possible bases are only found very rarely, we can also write this as *p_i,t_* for the SNP, with 1 − *p*_*i,t*_ the proportion of the reference base at that position at that time. We excluded all bases before position 55 and after position 29804 of the reference and ignored any deletions because these primarily create more noise in the data without adding signal.

## Proportion of SNPs over time

We obtain an initial estimate of fitness advantage for individual SNPs based on the increase (or decrease) of individual SNPs over time. This can be done by assuming that the SNP in question is the perfect representation of the variant; none of the wildtype viruses should contain the SNP while all of the variant viruses do contain the SNP. We further assume that both the wildtype virus population (*I*_*W*_) and the variant (*I*_*V*_) have a static growth rate, and consequently static basic reproduction numbers, unaltered by external factors such as build-up of immunity through new cases (i.e. the relation between their growth rates is fixed over time). We express the fitness advantage of the variant (*ρ*) as the difference in net growth rate between the wildtype and variant, similar to the approach used by others [4, 17],

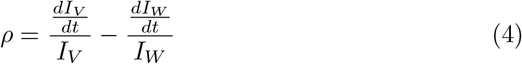

The proportion of infections caused by the variant among all infections thus follows a logistic growth curve,

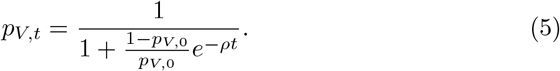

Note that the function of the proportion over time does not depend on the exact value of the growth rates or reproduction numbers of the wildtype and variant, but solely on the net difference between the growth rates.

## The logit of the variant proportion

We can convert the proportion of the variant over time to get to a more directly observable fitness advantage parameter by considering the logit of the proportion of a certain SNP (*p*_*s,t*_) instead of its untransformed proportion,

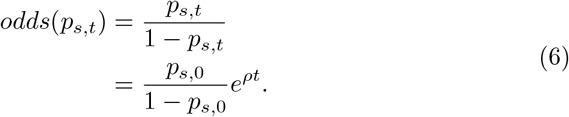

The logit (log odds) then gives us an easier form:

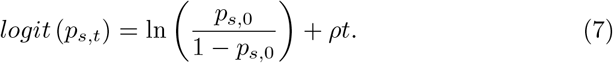

If we now consider the difference between the logit of the currently observed proportion (*t* = *t*) and the logit of the proportion at the (previously observed) start point (*t* = *t* −*τ*), we are left with just the term for the relative fitness advantage,

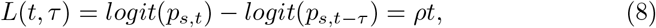

and then the advantage can be calculated as follows:

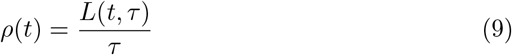

If we visualise the changes in proportions of all found SNPs between two consecutive weeks as a phase plane with the logit of the proportion in the previous week on the x-axis and the logit proportion in the current week on the y-axis, the perfect SNP should follow a straight line parallel to the diagonal at distance *L*(*t,* 7). At the same time, the original base in the wildtype population should decrease at the same distance (−*L*(*t,* 7)).

## Population genetic structure

The above method uses the measurements of single SNP proportions to estimate the fitness advantage of the variant based on the assumption that any SNP is either part of the wildtype or part of the variant. Instead, it is also possible to measure the change in genetic population structure over time to infer the likely fitness advantage of a variant underlying this change. We employ a method analogous to Wright’s F-Statistic (*F*_*ST*_), specifically adapted for whole genome sequence data of pathogen populations [6], to produce single estimates of fitness advantage of emerging variants for each week in the data.

In the predominant definition[18, 10, 9], *F*_*ST*_ measures the genetic divergence between populations by comparing the diversity (or heterozygocity) within the combined populations (*H*_*s*_) to the diversity between the populations (*H*_*p*_),

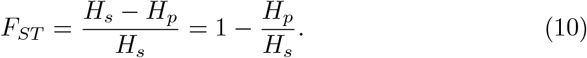

While *F*_*ST*_ is usually used to measure the divergence of contemporaneous populations in different locations, it can in principle also measure the amount the current population diverges from past populations in the same location. If the population is under strong selective pressure, or if more fit individuals are introduced, the genetic population structure can rapidly change. This change should be reflected in a *F* measurement between the current (*t* = 0) and a past or future (*t* + *τ*) population.

Since the logit-transformed proportions of variant-linked SNPs develop linearly over time, we get the logit-transformed *F_ST_*,

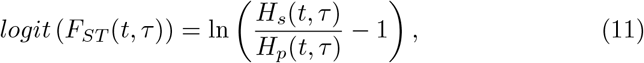

with a theoretical range of (− ∞,∞). To avoid extreme values for similar populations we instead used

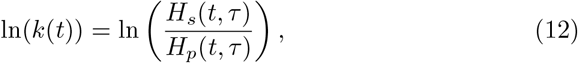

which ranges (0,∞) which defines the populations’ divergence as the diversity within the combined population relative to the diversity between the populations. As we compare populations from different time periods from the same location, we thus need to calculate the diversity within the combined population at the current (*t*) and a past or future (*t* + *τ*) time point (*H*_*s*_(*t, τ*)) as well as the diversity between both populations (*H*_*p*_(*t, τ*)).

We define diversity as the chance of picking the same discordant pair of bases at a given genomic position *i* from either population, summed over all positions, based on the proportion of sequences with this base at this position from the population at the given time point (*p*_*i,t*_),:

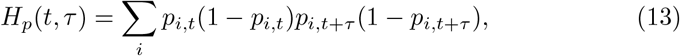

as well as the chance of picking the same discordant pair twice from the combined population,

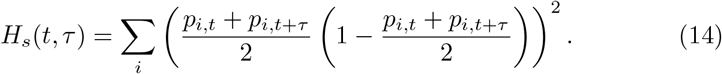

## Temporal population divergence

For simplicity, we will first consider the case where only one SNP is present in the population, removing the summation in both numerator and denominator, as well as subscript *i* in *p*_*i,t*_. Once a new variant starts spreading through the host population, it will increase its proportion over time following the equation in the previous section. The SNPs associated with the new variant will therefore start affecting the ln(*k*(*t*)) values, as their abundance starts to change as well. We can therefore describe the SNP proportion at future time point *t* + *τ* (*p*_*t*+*τ*_) as a function of the proportion at the current time (*p*_*t*_), given a particular fitness advantage *ρ*. Given the equation 8 in the previous section, we can express this advantage as the expected difference in the logit of the proportions, *L*(*t, τ*):

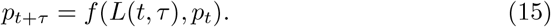

The exact form of this function can be derived by rewriting the definition of *L*(*t, τ*), as given by equation 8 as

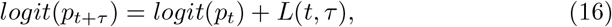

which means that

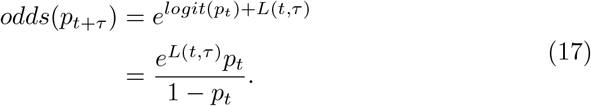

The expected proportion of the variant SNP therefore is a direct function of *p*_*t*_ and *L*(*t, τ*):

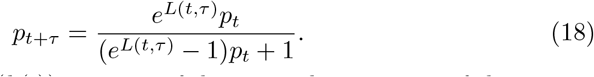

We can thus define ln(*k*(*t*)) in terms of the original proportion of the variant SNP (*p*_*t*_) and the difference in the logit of the proportion variant SNP in the population at both time points (*L*(*t, τ*)),

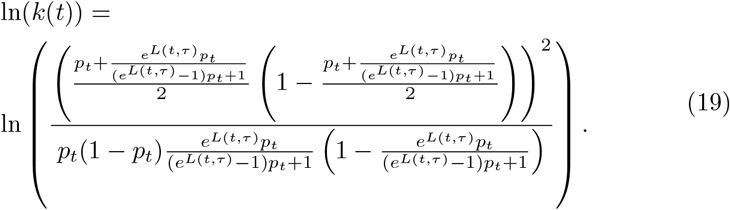

For larger absolute values of *L*(*t, τ*), thus with higher fitness advantage and/or longer time between observations, ln(*k*(*t*)) is approximately linearly related to *L*(*t, τ*) (See supplementary text section S7):

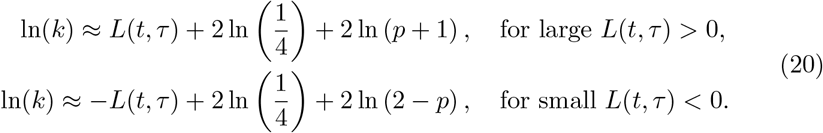

This slope translates directly to the fitness advantage *ρ* of the variant spreading within the sampled population.

Given that the above equation assumes a single SNP, we need to convert this to include all SNPs. This can be done by using the sum of all SNP-specific *H*_*s,i*_(*t, τ*) and *H*_*p,i*_(*t, τ*) values:

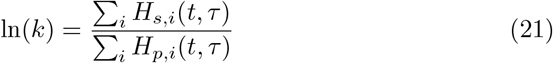

We then used the weeks leading up to the each reference week (2 ≥*τ*≥ 8) to estimate the fitness advantage of a potential variant growing at that moment in time. Subsequently, we designated any week with *ρ* ≥ 5.0% as having a potential variant spreading through the population (variant weeks). We then linked consecutive variant weeks into time periods under the assumption that their estimated fitness advantage was caused by the same underlying variant.

For reference, we show the percentage increase in transmissibility under the assumption of a wildtype effective reproduction number (*R*_*w*_) of 1, and a length of disease *D* of 5.2 days. Following [4], we then obtain the increase in transmissibility as

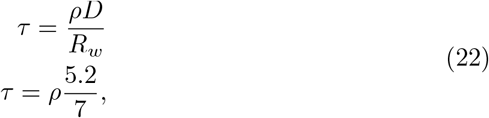

taking into account that *ρ* is measured per week.

## Sensitivity analysis

To assess the minimum number of sequences per week required to make a reliable estimate, we randomly sampled a given number of sequences from each week between 01-01-2021 and 01-10-2022, repeated a 100 times. This was done using 10, 25, 50, 100, 250, and 500 sequences. We subsequently assessed if the resulting periods of increased fitness as well as the maximum value of *ρ* overlapped with the original. In order to test the scalability of our algorithm, we also measured the time it took to perform each iteration. Furthermore, we repeated the analysis on datasets containing all sequences from the UK (N=2,775,418), the United States (N= 3,769,411), Denmark (N=576,296), and South Africa (N=35,770).

We also assessed the detection of increased fitness while taking the submission date of the isolates into account. To that end, we selected all isolates submitted before a given week, and estimated *ln*(*k*(*t*)) for all preceding weeks. Repeating this for all weeks in the dataset, we recorded the increase in *L*(*t,* 7) for the week before each reference week, giving a live view of the detection of increased fitness as newly submitted isolates are coming in.

## Live detection

To assess the performance of the method in real-time, we created sub-datasets for each reference week including only those isolates submitted up to (and including) the reference week. We then calculated the complete Δ ln(*k*) time line for each of these datasets, using their respective cut-off weeks as reference weeks. Subsequently, we also used one, two, and three weeks before as the reference week to account for the lack of data in the last weeks leading up to the cut-off week.

## Assigning isolates to the variant

For each of the time periods during which the spread of a variant with increased fitness was detected, we calculated per submitted sequence *j* the total SNP advantage score (*S*_*j*_). This SNP score is the sum of all differences in logit proportions at the start 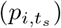 and end 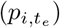 of the time period of all SNPs present in a particular sequence.

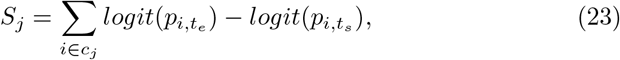

where *c*_*j*_ is the set of SNPs present in sequence *j*.

A positive score means that the sequence in question includes SNPs that generally increase over the time period, and is therefore likely part of the variant population, while negative scores mean it is likely part of the wild-type population. We then compared for each time period the sequence scores with the general grouping of Pango-lineages (Table S2) to test the agreement of the SNP scoring with the general consensus on variant classification.

## Data, Materials, and Software Availability

All used data are available via the GISAID Initiative. R code for the analysis is available at https://github.com/QUPI-IUK/fitnessGainEstimate

## Acknowledgments

This publication was funded by the German Federal Ministry of Education and Research (BMBF) Network of University Medicine 2.0: NUM 2.0”, Grant No. 01KX2121, Project: Genomic Pathogen Surveillance and Translational Research (GenSurv). We thank Fabian Brkin for useful discussions on measuring the method’s performance. We gratefully acknowledge all data contributors, i.e., the Authors and their Originating laboratories responsible for obtaining the specimens, and their Submitting laboratories for generating the genetic sequence and metadata and sharing via the GISAID Initiative, on which this research is based.

