## Supplementary information for "Estimation of SARS-CoV-2 fitness gains from genomic surveillance data without prior lineage classification"

#### Section S1 Supplementary figures

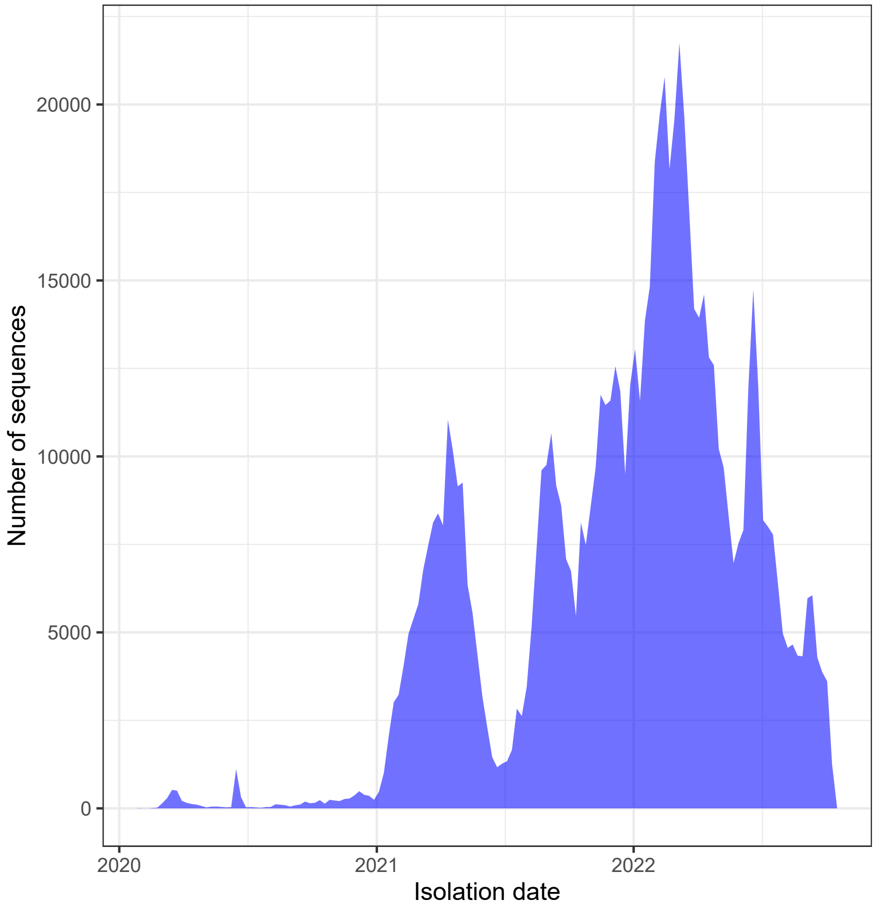


Figure S1: The number of collected isolates from Germany in the GISAID data, by isolation date.

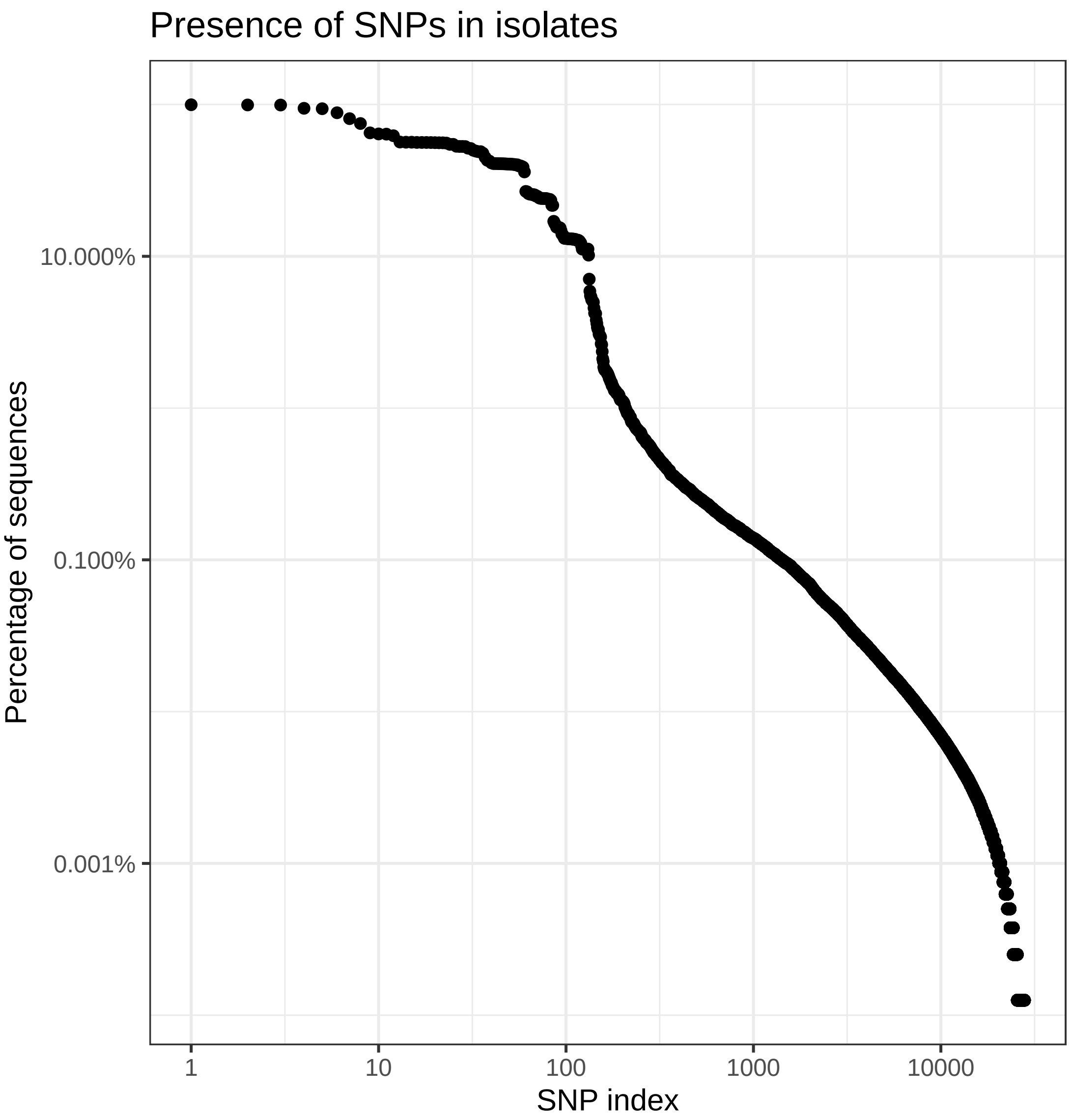


Figure S2: The proportion of isolates containing a particular SNP, on a log-log scale. SNPs are ordered by their proportion presence.

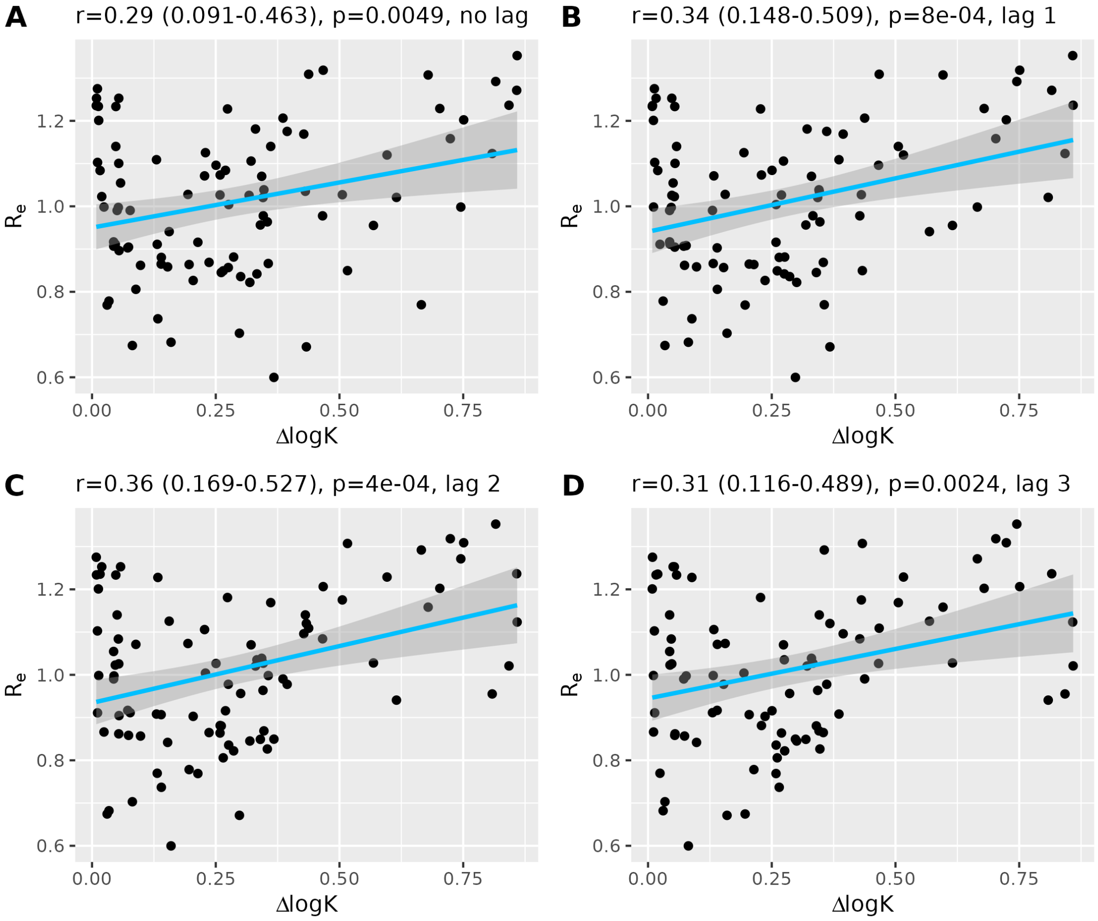


Figure S3: The correlation between the measured $\Delta \ln(k)$ and the time-varying reproduction number $R_e$ based on the number of reported cases in Germany. Each panel shows a different lag between $\ln(k)$ and $R_e$, to account for a delayed effect of the fitness gain on the reproduction number. A two week lag delivers the best correlation, indicating that the $R_e$ tends to increase after $\Delta \ln(k)$ increases.

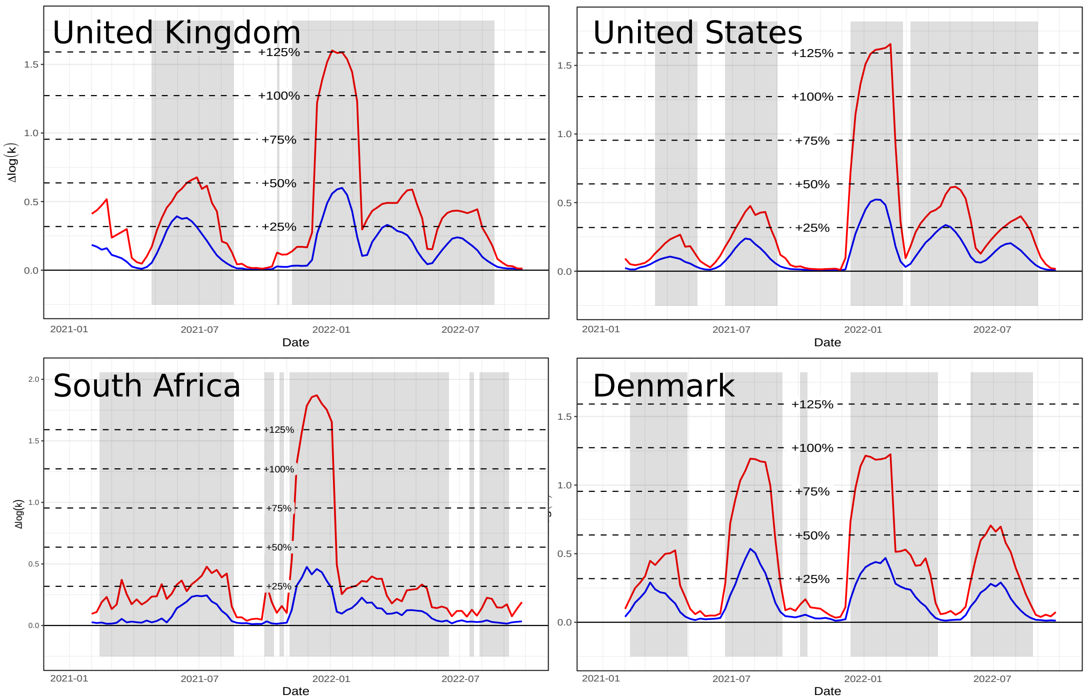


Figure S4: Estimates of fitness gain over time for other countries, showing the UK, US, South Africa, and Denmark.

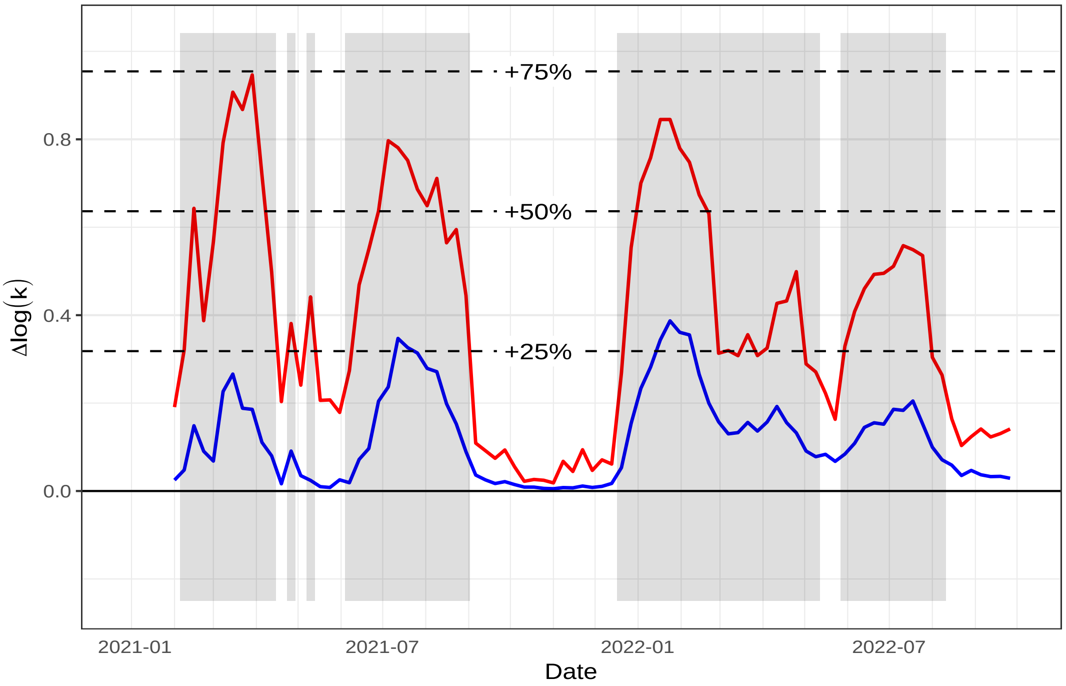


Figure S5: Estimates of fitness gain over time for the state of Baden-Württemberg.

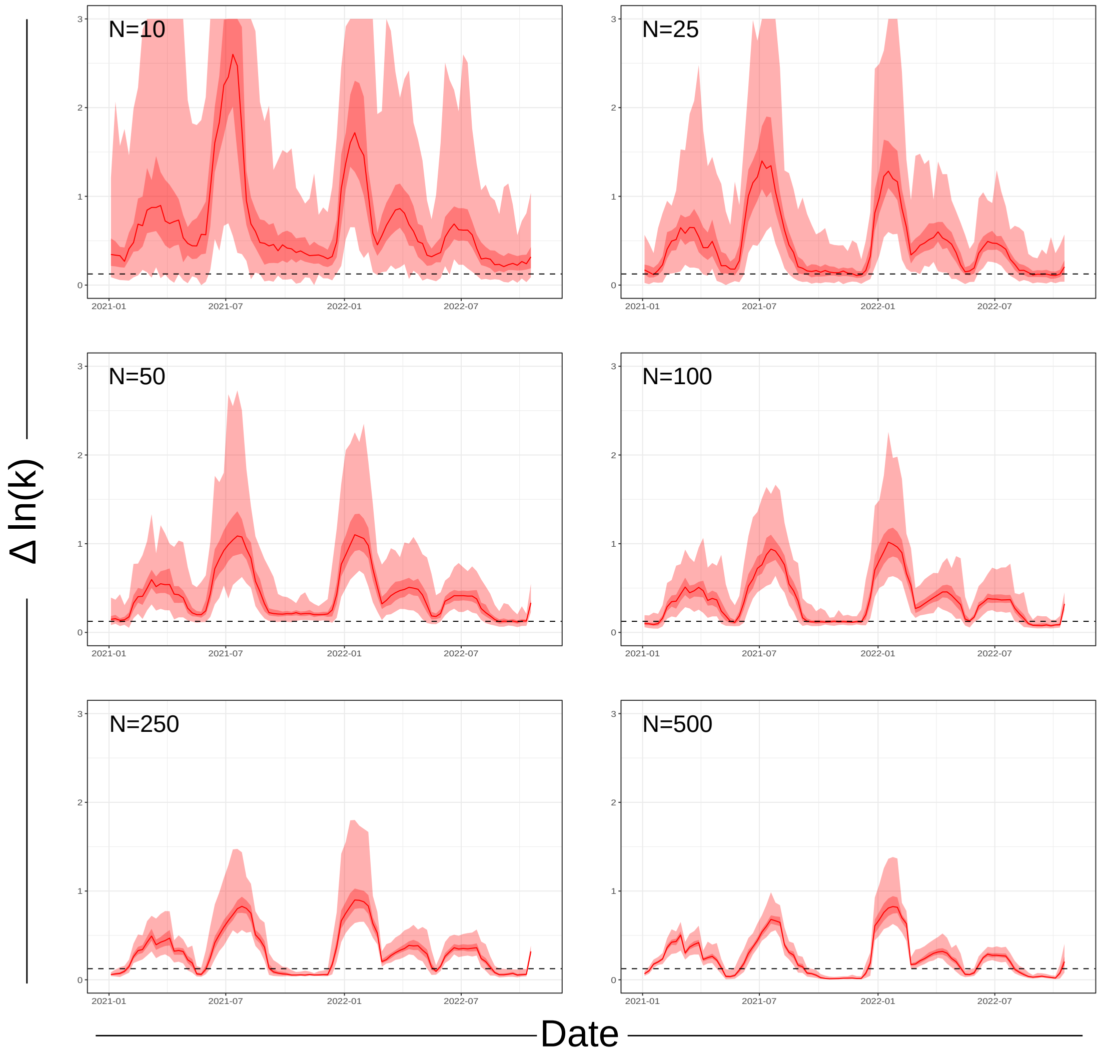


Figure S6: The influence of the size of the number of isolates included each week. We resampled 10,25,50,100, 250, and 500 isolates from each week and calculated the $\Delta \ln(k)$ value, repeated 100 times. Light shaded areas show the full range, dark range the interquartile range, and the solid line the median.

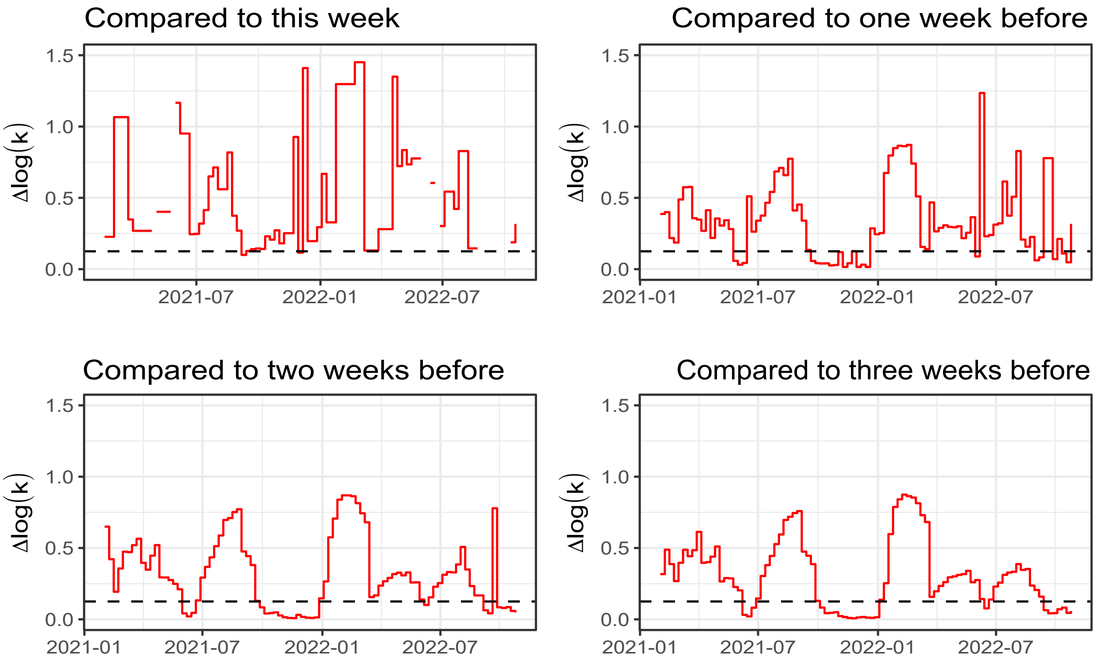


Figure S7: Estimates of fitness gain based on submission dates of the isolates. For each week, we calculated $\Delta \ln(k)$ based on the samples submitted until that week. We than used the submission week as the focal week (used for the pairwise comparison of genetic population structure), as well as the three weeks before. This is done because the submission week often contains too few isolates to make a proper comparison.

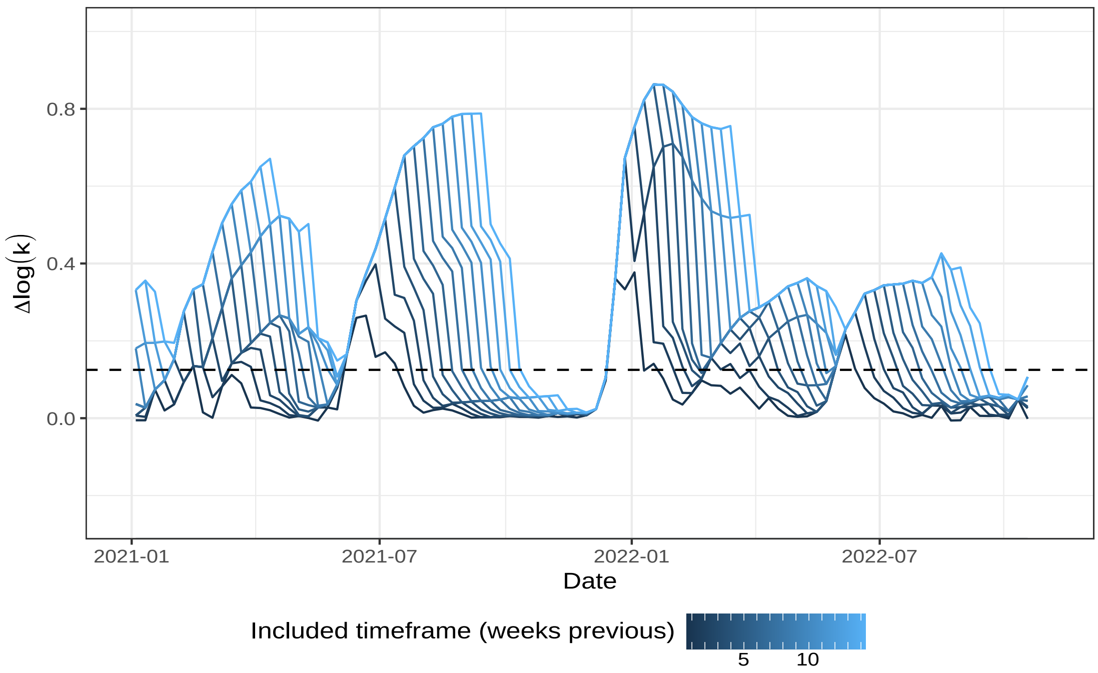


Figure S8: The influence of the size of the timeframe being compared to the reference week. We incrementally increased the number of days in the past included in the $\Delta \ln(k)$ calculation. Larger timeframes generally result in higher estimates, but rarely result in earlier detection.

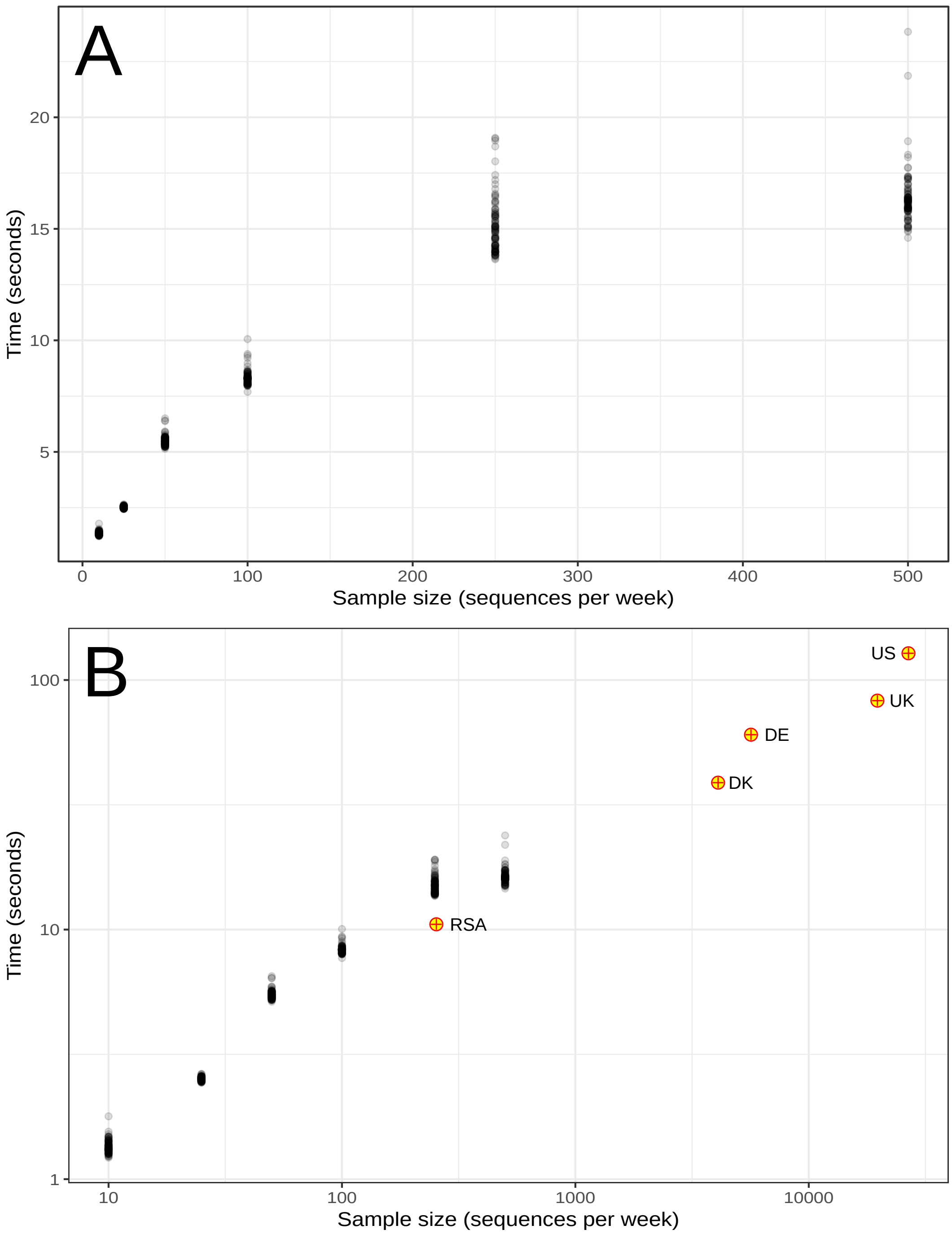


Figure S9: The time required to produce an estimate as a function of the number of included isolates per week. (A) For all the resampling repeats, (B) including the results for the different countries, on a log-log scale. Note that for the countries, the number of sequences per week is the total number of sequences divided by the total number of weeks in the data.

### Section S2 GISAID data

#### Data Availability

| | |
|---|---|
| GISAID Identifier: | EPI_SET_230724bm |
| DOI: | 10.55876/gis8.230724bm |

All genome sequences and associated metadata in this dataset are published in GISAIDs EpiCoV database. To view the contributors of each individual sequence with details such as accession number, Virus name, Collection date, Originating Lab and Submitting Lab and the list of Authors, visit 10.55876/gis8.230724bm .

#### Data Snapshot

- EPI_SET_230724bm is composed of 148,410 individual genome sequences.
- The collection dates range from 2020-02-25 to 2023-10-11;
- Data were collected in 1 countries and territories;
- All sequences in this dataset are compared relative to hCoV-19/Wuhan/WIV04/2019 (WIV04), the official reference sequence employed by GISAID (EPI_ISL_402124). Learn more at https://gisaid.org/WIV04.

#### Section S3 SNP variation

The main methods assume that the analysed SNP is a perfect representation of the variant in question, with its particular fitness advantage over the background population. This assumption, of course, doesn't hold in actual pathogen populations, due to genetic variation through random drift. In this context, it is important to consider that not all SNPs found in the variant attribute to the fitness advantage, i.e. are positively selected for. Quite a number of neutral (or near-neutral) SNPs may hitch-hike with the SNPS conferring the fitness advantage. However, neutral SNPs would not arise at that exact same moment as the "perfect" variant SNPs, and their trajectories would therefore differ, depending on the time they arose relative to the emergence of the variant. Those neutral SNPs that arose before the variant will be shared between the entire variant and part of the wildtype population (wildtype variation), while those neutral SNPs that arose in the variant population after the positive SNPs arose should be present in the entire wildtype population and only part of the variant population (variant variation).

Neutral SNPs arising in the wildtype population after the variant arose fall within the wildtype variation, but they will have a very limited effect since the relative abundance of the wildtype population declines from the time the variant arose. We consider the probability of homoplasy occurring too low to be incorporated in the fitness advantage estimations. This primarily because wildtype and variant variation are considered to be te result of neutral mutation, with the probability of the same random mutation happening twice sufficiently small to be disregarded. We therefore assume that none of the SNPs are partially present in both the variant and the wildtype population.

In general, the SNP proprtion can be described as follows:

$$p_{s,t} = \alpha p_{v,t} + \beta(1 - p_{v,t}), \tag{1}$$

where $\alpha$ denotes the proportion of the variant population with the SNP present, and $\beta$ the proportion of the wildtype population with the SNP present. Because we assume that homoplasy is rare, we can set $\beta$ to 0 or 1 and allow $\alpha$ to be anything between 0 or 1 (i.e. the SNP is part of variation in the VoC), or set $\alpha$ to 0 or 1 and allow $\beta$ to vary. This means that we are left with only a four combinations

| | | | |
|---|---|---|---|
| 1) | $\alpha = \frac{p_{s,t}}{p_{v,t}}$ | $\beta = 0$ | $p_{s,t} \leq p_{v,t}$ |
| 2) | $\alpha = \frac{p_{s,t}-(1-p_{v,t})}{p_{v,t}}$ | $\beta = 1$ | $p_{s,t} \geq 1 - p_{v,t}$ |
| 3) | $\alpha = 0$ | $\beta = \frac{p_{s,t}}{1-p_{v,t}}$ | $p_{s,t} \leq 1 - p_{v,t}$ |
| 4) | $\alpha = 1$ | $\beta = \frac{p_{s,t}-p_{v,t}}{1-p_{v,t}}$ | $p_{s,t} \geq p_{v,t}$ |

Table S1: Rules for variation in the wildtype and variant population

Combinations 1 and 4 concern SNPs that will increase in frequency over time, while the SNPs from combinations 2 and 3 decrease in frequency, given that we define the variant as having a positive fitness advantage.

Neutral SNPs in the wildtype will appear to increase slower in frequency than the actual variant, as they move from its proportion within the wildtype population, which is initially higher than the variant, towards the variant proportion. Similarly, neutral SNPs in the variant will also appear to increase slower, as the final proportion of their trajectory is their proportion within the variant, which is lower than the final proportion of the variant's trajectory (i.e., 1).

Given equation 1, we expect most SNPs to be present on one of two straight lines on the untransformed proportion phase plane: either on the line connecting $(p_{v,t}, p_{v,(t+1)})$ with $(0, 0)$, or on the line connecting $(p_{v,t}, p_{v,(t+1)})$ with $(1, 1)$. SNPs present on the first line are partially present in the variant population, while SNPs present on the second line are partially present in th wildtype population and fully present in the variant population.

If we assume that the observed SNPs with a significant fitness advantage are the result of a single variant growing in the viral population, we need to find a way to join this with the observation that not all SNPs are present at the same proportion at the same times. We estimated the fitness of the variant for each week by fitting equation 1 to the SNP proportions in both the current and the previous week, using the rules tabulated in table S1.

This method is able to estimate the fitness advantage as well as the current proportion of the population containing the advantage for weeks when the signal is strong (i.e. when proportions deviate significantly from 0 or 1). However, when the variant proportion is low, or no variant is present, it performs poorly, often estimating high fitness advantages for low prevalence variant (figure S10C).

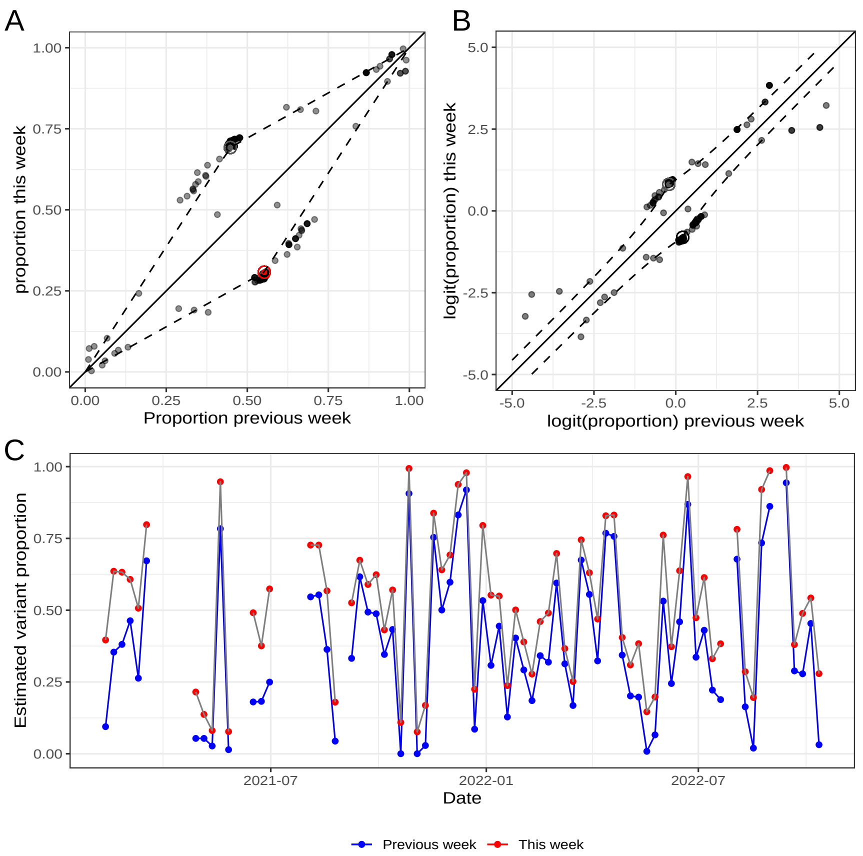


Figure S10: Estimating variant fitness advantage. A) Any SNP proportions (gray dots) deviating from the variant and wild-type proportions (red dots) are assumed to be caused by variation within these populations and should follow the dashed lines. B) Similar but based on the logit of the proportions. C) The estimated variant proportion in each week, and its preceding week, with sufficiently included SNPs.

#### Section S4 Additional information on the post-hoc classification of variants

| Variant | Included Pango lineages |
|---|---|
| Alpha | B.1.1.7<br>B.1.1.7.* |
| Delta | B.1.617.2<br>B.1.617.2.*<br>AY.* |
| Omicron BA.1 | B.1.1.529<br>B.1.1.529.*<br>BA.1<br>BA.1.* |
| Omicron BA.2 | BA.2<br>BA.2.* |
| Omicron BA.3 | BA.3<br>BA.3.* |
| Omicron BA.2 | BA.5<br>BA.5.*<br>BE.*<br>BF.* |

Table S2: Grouping of Pango lineages into variant groups.

#### Section S5 Isolate SNP scores

| | Negative | Positive | Total |
|---|---|---|---|
| alpha | 7119 | 87733 | 94852 |
| Other | 8678 | 7080 | 15758 |
| Total | 15797 | 94813 | 110610 |

Table S3: Total SNP score of isolates between 14-02-2021 and 03-04-2021 by lineage (Alpha or other). Sensitivity: 92.81%, specificity: 75.06%.

| | Negative | Positive | Total |
|---|---|---|---|
| Delta | 4 | 56839 | 56843 |
| Other | 6925 | 35 | 6960 |
| Total | 6929 | 56874 | 63803 |

Table S4: Total SNP score of isolates between 20-06-2021 and 21-08-2021 by lineage (Delta or other). Sensitivity: 99.99%, specificity: 99.5%.

| | Negative | Positive | Total |
|---|---|---|---|
| Omicron BA.2 | 0 | 183296 | 183296 |
| Other | 34640 | 119298 | 153938 |
| Total | 34640 | 302594 | 337234 |

Table S5: Total SNP score of isolates between 26-12-2021 and 14-05-2022 by lineage (BA.2 or other). Sensitivity: 100%, specificity: 22.5%.

| | Negative | Positive | Total |
|---|---|---|---|
| Omicron BA.1 | 74 | 117074 | 117148 |
| Omicron BA.2 | 0 | 183296 | 183296 |
| Other | 34566 | 2224 | 36790 |
| Total | 34640 | 302594 | 337234 |

Table S6: Total SNP score of isolates between 26-12-2021 and 14-05-2022 by lineage (BA.1, BA.2, or other). Sensitivity: 99.98%, specificity: 93.95%.

| | Negative | Positive | Total |
|---|---|---|---|
| Omicron BA.5 | 0 | 74537 | 74537 |
| Other | 13587 | 15022 | 28609 |
| Total | 13587 | 89559 | 103146 |

Table S7: Total SNP score of isolates between 05-06-2022 and 30-07-2022 by lineage (BA.5 or other). Sensitivity: 100%, specificity: 47.49%.

#### Section S6 Benchmark desktop computer

The configuration of our benchmark computer is as follows:

| Model | DELL Inc. OptiPlex 7060 (085A) |
| --- | --- |
| Processor | Intel(R) Core(TM) i7-8700 CPU @ 3.20GHz (6 cores, 12 threads) |
| Memory | Hynix Semiconductor (Hyundai Electronics) |
| | 64GiB (4x16GiB) DIMM DDR4 Synchronous 2666 MHz (0,4 ns) |

#### Section S7 Derivation and asymptotics

For $k = \frac{H_s}{H_p}$, we derive the logarithmic equations that govern the fitness advantage for a given proportion $p$.

We define the abbreviation $b$ as

$$b := \frac{e^{L(\tau)}p}{(e^{L(\tau)}-1)p+1},$$

and then compute

$$\begin{aligned}
\log(k) &= \log\left(\frac{H_s}{H_p}\right) \\
&= \log(H_s) - \log(H_p) \\
&= \log\left(\left(\frac{1}{4}(p+b)(2-(p+b))\right)^2\right) - \log(p(1-p)b(1-b)) \\
&= 2\log\left(\frac{1}{4}\right) + 2\log\left(p + \frac{e^{L(\tau)}}{(e^{L(\tau)}-1)p+1}\right) + 2\log\left(2 - \left(p + \frac{e^{L(\tau)}}{(e^{L(\tau)}-1)p+1}\right)\right) \\
&\quad - \log(p) - \log(1-p) - \log(b) - \log(1-b) \\
&= 2\log\left(\frac{1}{4}\right) + 2\log\left(e^{L(\tau)}(p+1) - (p-1)\right) - 2\log\left((e^{L(\tau)}-1)p+1\right) + 2\log(p) \\
&\quad + 2\log(2-p) + 2\log\left((e^{L(\tau)}-1)p + 1 - \frac{e^{L(\tau)}p}{(2-p)}\right) - 2\log\left((e^{L(\tau)}-1)p+1\right) \\
&\quad - \log(p) - \log(1-p) - \log\left(e^{L(\tau)}\right) - \log(p) \\
&\quad + \log\left((e^{L(\tau)}-1)p+1\right) - \log(1-p) + \log\left((e^{L(\tau)}-1)p+1\right). \qquad (2)
\end{aligned}$$

By using the identity $\log\left((e^{L(\tau)}-1)p+1\right) = \log\left(e^{L(\tau)}\right) + \log\left(p + \frac{1-p}{e^{L(\tau)}}\right)$ we can identify the asymptotic behaviour of $\log(k)$ when $L \to +\infty$. In the same manner we have $\log\left((e^{L(\tau)}-1)p + 1 - \frac{e^{L(\tau)}p}{(2-p)}\right) = \log\left(e^{L(\tau)}\right) + \log\left(p + \frac{1-p}{e^{L(\tau)}} - \frac{p}{2-p}\right)$ and analogously we find $\log\left(e^{L(\tau)}(p+1) - (p-1)\right) = \log\left(e^{L(\tau)}\right) + \log\left((p+1) - \frac{p-1}{e^{L(\tau)}}\right)$. Inserting these substitutions, the terms above then take the form

$$
\begin{aligned}
\log(k) = {} & 2\log\left(\frac{1}{4}\right) + 2\log\left(e^{L(\tau)}(p+1)-(p-1)\right) - 2\log\left(e^{L(\tau)}\right) - 2\log\left(p+\frac{1-p}{e^{L(\tau)}}\right) + 2\log(p) \\
& + 2\log(2-p) + 2\log\left((e^{L(\tau)}-1)p + 1 - \frac{e^{L(\tau)}p}{(2-p)}\right) - 2\log\left(e^{L(\tau)}\right) - 2\log\left(p+\frac{1-p}{e^{L(\tau)}}\right) \\
& - \log(p) - \log(1-p) - \log\left(e^{L(\tau)}\right) - \log(p) \\
& + \log\left(e^{L(\tau)}\right) + \log\left(p+\frac{1-p}{e^{L(\tau)}}\right) - \log(1-p) + \log\left(e^{L(\tau)}\right) + \log\left(p+\frac{1-p}{e^{L(\tau)}}\right) \\
= {} & 2\log\left(\frac{1}{4}\right) + 2\log\left(e^{L(\tau)}\right) + 2\log\left((p+1)-\frac{p-1}{e^{L(\tau)}}\right) - 2\log\left(e^{L(\tau)}\right) \\
& - 2\log\left(p+\frac{1-p}{e^{L(\tau)}}\right) + 2\log(p) \\
& + 2\log(2-p) + 2\log\left(e^{L(\tau)}\right) + 2\log\left(p+\frac{1-p}{e^{L(\tau)}}-\frac{p}{2-p}\right) \\
& - 2\log\left(e^{L(\tau)}\right) - 2\log\left(p+\frac{1-p}{e^{L(\tau)}}\right) \\
& - \log(p) - \log(1-p) - \log\left(e^{L(\tau)}\right) - \log(p) \\
& + \log\left(e^{L(\tau)}\right) + \log\left(p+\frac{1-p}{e^{L(\tau)}}\right) - \log(1-p) + \log\left(e^{L(\tau)}\right) + \log\left(p+\frac{1-p}{e^{L(\tau)}}\right) \\
= {} & \log\left(e^{L(\tau)}\right) + 2\log\left(\frac{1}{4}\right) - 2\log(1-p) + 2\log(2-p) \\
& - 2\log\left(p+\frac{1-p}{e^{L(\tau)}}\right) + 2\log\left((p+1)-\frac{p-1}{e^{L(\tau)}}\right) \\
& + 2\log\left(p+\frac{1-p}{e^{L(\tau)}}-\frac{p}{2-p}\right).
\end{aligned}
$$

The asymptotic equation for large $L$ is then given by the following scaling law.

$$
\begin{aligned}
\log(k) \approx {} & L + 2\log\left(\frac{1}{4}\right) - 2\log(1-p) + 2\log(2-p) \\
& - 2\log(p) + 2\log(p+1) \\
& + 2\log\left(p-\frac{p}{2-p}\right) \\
= {} & L + 2\log\left(\frac{1}{4}\right) + 2\log(p+1).
\end{aligned}
$$

From the equations that we derived just before we substituted some expressions, namely (2), we find the asymptotic behaviour of $\log(k)$ for very small $L$ by noting that $e^{L(\tau)} \to 0$ for

$L \to -\infty$. The final approximation for small enough $L < 0$ then reads

$$\begin{aligned}
\log(k) \approx\ & 2\log\left(\frac{1}{4}\right) + 2\log(1-p) - 2\log(1-p) + 2\log(p) \\
& + 2\log(2-p) + 2\log(1-p) - 2\log(1-p) \\
& - \log(p) - \log(1-p) - L - \log(p) \\
& + \log(1-p) - \log(1-p) + \log(1-p) \\
=\ & -L + 2\log\left(\frac{1}{4}\right) + 2\log(2-p)\,.
\end{aligned}$$

Both approximations for $\log(k)$ then can be transformed by an exponential to get back simple expressions for $k$ when the absolute value of $L$ is large.

$$\begin{aligned}
k &\approx e^{L}\left(\frac{1}{4}\right)^{2}(p+1)^{2}\,, \qquad \text{for large } L > 0, \\
k &\approx e^{-L}\left(\frac{1}{4}\right)^{2}(2-p)^{2}\,, \qquad \text{for small } L < 0.
\end{aligned}$$

Since we are also interested in the slope, we computed the derivative of the exact equation with respect to $L$ as well.

$$\begin{aligned}
\frac{\partial}{\partial L}\log(k) =\ & \frac{e^{L(\tau)}}{e^{L(\tau)}} + 2\frac{\frac{1-p}{e^{L(\tau)}}}{p + \frac{1-p}{e^{L(\tau)}}} - 2\frac{\frac{p-1}{e^{L(\tau)}}}{(p+1) - \frac{p-1}{e^{L(\tau)}}} \\
& - 2\frac{\frac{1-p}{e^{L(\tau)}}}{\left(p + \frac{1-p}{e^{L(\tau)}} - \frac{p}{2-p}\right)} \\
=\ & 1 + \frac{2}{e^{L(\tau)}}\left(\frac{1-p}{p + \frac{1-p}{e^{L(\tau)}}} - \frac{p-1}{(p+1) - \frac{p-1}{e^{L(\tau)}}} - \frac{1-p}{\left(p + \frac{1-p}{e^{L(\tau)}} - \frac{p}{2-p}\right)}\right).
\end{aligned}$$

In the limit $L \to \infty$, the slope thus equals 1.
